# Prevalence and Determinants of Teenage Pregnancy in Cambodia: A Multilevel Analysis of the 2021–22 Demographic and Health Survey

**DOI:** 10.64898/2025.12.09.25341934

**Authors:** Yem Sokha, Kem Sokunthea, Tun Sreypeov

## Abstract

**Background:** Teenage pregnancy remains a significant public health challenge in Cambodia, with implications for maternal and child health outcomes, educational attainment, and socioeconomic development.

**Objective:** To determine the prevalence of teenage pregnancy among Cambodian women aged 15-19 years and identify its individual and community-level determinants using multilevel analysis of data from the 2021-22 Cambodia Demographic and Health Survey (CDHS).

**Methods:** We analyzed data from 5,783 adolescent women aged 15-19 years from the nationally representative CDHS 2021-22. Given the hierarchical structure of DHS data (individuals nested within 709 clusters), we employed multilevel logistic regression models. Four nested models were fitted, including a null model with random intercept only, an individual-level model including age, education, marital status, wealth, contraceptive use, and internet use, a community-level model including residence and distance to health facilities, and a full model combining both levels. Intraclass correlation coefficient (ICC) and likelihood ratio tests assessed clustering effects. We reported adjusted odds ratios (AOR) with 95% confidence intervals.

**Results:** The weighted prevalence of teenage pregnancy was 28.08% (95% CI: 26.48-29.75). The ICC indicated 10.07% of variance in teenage pregnancy was attributable to cluster-level factors, strongly justifying multilevel modeling (LR test comparing multilevel to standard logistic regression: χ²=52.69, df=1, p<0.001). In the full multilevel model, older adolescent age (18-19 vs 15-17 years: AOR=2.73, 95% CI: 1.80-4.14, p<0.001), current marriage (AOR=4,611.27, 95% CI: 1,929.44-11,020.73, p<0.001), and former marriage (AOR=434.84, 95% CI: 132.85-1,423.27, p<0.001) were significantly associated with increased odds of teenage pregnancy. Higher education showed protective effects (AOR=0.06, 95% CI: 0.01-0.60, p=0.017). Wealthier households demonstrated lower odds (richest vs poorest: AOR=0.22, 95% CI: 0.11-0.46, p<0.001). Current contraceptive use was associated with higher odds (AOR=4.69, 95% CI: 2.85-7.71, p<0.001), reflecting post-conception initiation rather than primary prevention. After accounting for individual-level factors, community-level variables (rural residence, distance to facilities) were not independently significant.

**Conclusions:** More than one in four Cambodian adolescent girls experience teenage pregnancy, with substantial between-cluster variation (ICC=10.07%). Early marriage is the dominant pathway to teenage pregnancy. Multilevel interventions addressing child marriage prevention, educational retention, socioeconomic empowerment, and youth-friendly reproductive health services are urgently needed.

## INTRODUCTION

### Background and Significance

Teenage pregnancy remains a pressing public health challenge globally, affecting approximately 21 million girls aged 15-19 years annually in developing regions (United Nations Population Fund [UNFPA], 2023). In Southeast Asia, adolescent pregnancy rates vary substantially across countries (Raj et al., 2020; World Health Organization [WHO], 2021), with Cambodia experiencing persistent challenges despite socioeconomic development and health system improvements over recent decades (National Institute of Statistics [NIS] & ICF, 2022). Teenage pregnancy carries significant consequences for both adolescent mothers and their children. Health risks include increased maternal mortality, obstetric complications, preterm delivery, and low birth weight (Ganchimeg et al., 2014; WHO, 2020). Beyond health outcomes, teenage pregnancy perpetuates cycles of poverty through educational interruption, limited employment opportunities, and reduced lifetime earnings (Azevedo et al., 2012; Psaki et al., 2022). Children born to adolescent mothers face elevated risks of developmental delays, malnutrition, and mortality (Fall et al., 2015; Finlay et al., 2017).

### The Cambodian Context

Cambodia has made remarkable progress in maternal and child health over the past two decades, with substantial reductions in maternal and under-five mortality rates (NIS & ICF, 2022; World Bank, 2023). However, adolescent reproductive health remains a concern. The 2014 Cambodia Demographic and Health Survey (CDHS) reported that 12% of women aged 15-19 had begun childbearing, highlighting teenage pregnancy as a priority public health issue (NIS, Directorate General for Health [DGH], & ICF International, 2015). The prevalence of pregnancy among women aged 15-19 varies significantly by multiple intersecting factors. Age demonstrates a steep gradient, with older teenagers (18-19 years) showing substantially higher rates than younger adolescents (NIS et al., 2015). Educational attainment exhibits a strong inverse association, with pregnancy rates declining progressively from no education through higher education (Lloyd & Mensch, 2008). Marital status shows the starkest division: nearly all married adolescents experience pregnancy, while rates among never-married adolescents remain very low (Godha et al., 2013). Socioeconomic conditions, particularly household wealth, demonstrate consistent protective gradients, with poorer households facing substantially elevated risk (Raj et al., 2020). Early marriage and sexual debut, often driven by cultural and family expectations, are key contributors to adolescent pregnancy (Parsons et al., 2015; Raj et al., 2009; United Nations Children’s Fund [UNICEF], 2021). Additionally, limited contraceptive knowledge, use, and unmet family planning needs heighten vulnerability among teenagers (Chandra-Mouli et al., 2014; WHO, 2020).

### Methodological Gap

Despite recognition of teenage pregnancy as a public health priority, comprehensive, nationally representative analyses using recent data and appropriate statistical methods are limited (Blanc et al., 2013). Previous studies have not utilized multilevel modeling to address the hierarchical structure inherent in DHS data where individuals are nested within clusters, which are nested within provinces (Merlo et al., 2006; Rodriguez & Goldman, 2001). This methodological gap can lead to underestimated standard errors, inflated Type I error rates, and inability to quantify contextual effects (Heeringa et al., 2017; Snijders & Bosker, 2012).

### Theoretical Framework

This study is guided by socioecological theory (Bronfenbrenner, 1979), which posits that adolescent behaviors and outcomes result from interactions across multiple levels including individual factors such as age and education, interpersonal factors such as marital relationships and family dynamics, community factors such as local norms and service availability, and societal factors such as policies and cultural beliefs. Our multilevel analytical approach operationalizes this framework by explicitly modeling individual and community levels, allowing us to distinguish compositional effects representing individual characteristics clustered geographically from contextual effects representing place-based influences on similarly situated individuals. This framework has proven valuable in understanding adolescent reproductive health in low- and middle-income countries (Magadi & Agwanda, 2010; Stephenson et al., 2007).

### Study Rationale and Contribution

The 2021-22 CDHS provides an opportunity to assess current prevalence and identify determinants using rigorous multilevel analytical approaches (NIS & ICF, 2022). This study contributes to the literature by employing multilevel logistic regression to properly account for clustering effects (Merlo et al., 2006; Snijders & Bosker, 2012), quantifying between-cluster variation through intraclass correlation coefficient (ICC) and random effects, distinguishing individual and community-level determinants, using recent post-pandemic data to inform current policy, and following internationally recognized reporting standards for multilevel DHS analyses (Heeringa et al., 2017).

### Objectives

The primary objective is to estimate the prevalence of teenage pregnancy among women aged 15-19 years in Cambodia using data from the 2021-22 CDHS. The secondary objectives are to quantify between-cluster variation in teenage pregnancy using multilevel modeling, to identify individual sociodemographic determinants including age, education, marital status, wealth, and contraceptive use, to examine community-level characteristics including urban-rural residence and distance to health facility, and to determine which factors remain independently associated with teenage pregnancy after multilevel adjustment.

## METHODS

### Study Design and Data Source

We conducted a secondary data analysis using the nationally representative Cambodia Demographic and Health Survey (CDHS) 2021-22 dataset (NIS & ICF, 2022, 2023). The CDHS is a cross-sectional, population-based survey conducted as part of the global DHS Program (Corsi et al., 2012), collecting data on demographic characteristics, reproductive health, maternal and child health, and related indicators.

### Sampling Design

The CDHS 2021-22 employed a two-stage stratified cluster sampling design to ensure representativeness across urban and rural areas and all 25 provinces of Cambodia (NIS & ICF, 2022). In the first stage, 709 enumeration areas (clusters) were selected with probability proportional to size from a sampling frame based on the 2019 Cambodia General Population Census. In the second stage, systematic random sampling was used to select approximately 28 households per cluster. The survey achieved a response rate of 96.2% for the women’s questionnaire (NIS & ICF, 2022), which exceeds the recommended threshold for population-based surveys (Johnson & Wislar, 2012).

### Study Population

The study population comprised all women aged 15-19 years who participated in the CDHS 2021-22 women’s questionnaire (N = 5,783). This age range is internationally recognized as the teenage or adolescent period where pregnancy poses significant health and social risks (WHO, 2020). We included all adolescents regardless of marital status.

### Sample Size and Statistical Power

The achieved sample of 5,783 adolescents provides adequate statistical power for our analyses. With a prevalence of 28% and design effect of 1.98, the effective sample size of approximately 2,920 yields 80% power to detect odds ratios of 1.5 or greater for exposures with 20% prevalence at α=0.05 (Fleiss et al., 2003). For multilevel modeling, Maas and Hox (2005) recommend at least 50 clusters with 30 observations per cluster, and our 709 clusters with mean cluster size of 8 adolescents provides adequate power for detecting cluster-level variance and estimating random effects reliably.

### Variables and Measurements

We operationalized teenage pregnancy status as a binary variable indicating whether an adolescent had ever been pregnant or had a live birth. This outcome was constructed using two CDHS variables: total children ever born (v201), coded as positive if greater than zero, and current pregnancy status (v213), coded as positive if the respondent was pregnant at the time of the survey. Adolescents meeting either criterion were classified as having experienced teenage pregnancy. This operationalization aligns with standard DHS approaches to measuring adolescent fertility (Croft et al., 2018; NIS & ICF, 2022) and follows WHO definitions of adolescent pregnancy (WHO, 2020).

Following the hierarchical structure of DHS data, we categorized independent variables into two levels to reflect the nested nature of individuals within communities (Heeringa et al., 2017). Level 1 variables captured individual-level factors, including age group comparing 15 to 17 years with 18 to 19 years (v013), education level categorized as no education, primary, secondary, or higher (v106), marital status (v501), household wealth quintile (v190), current contraceptive use coded as yes or no (v312), and internet use indicating whether the respondent had ever used internet coded as yes or no (v171a). For marital status, we classified adolescents as never married, currently married or cohabiting, or formerly married including those who were widowed, divorced, or separated, with the original v501 categories such as “not living together” and “no longer living together or separated” collapsed into the formerly married group to ensure adequate cell sizes for analysis. The wealth index used in the CDHS is derived from principal components analysis of household assets and living conditions and is reported in five quintiles from poorest to richest (NIS & ICF, 2022; Rutstein & Johnson, 2004). Level 2 variables represented community-level characteristics, specifically place of residence comparing urban with rural areas (v025) and perceived distance to a health facility as a big problem versus not a big problem (v467d). The primary sampling unit or cluster (v021) was used as the grouping variable for the multilevel models.

### Statistical Analysis

We conducted all analyses using Stata version 17.0 (StataCorp, 2021), accounting for the complex survey design using survey weights (v005/1,000,000), clustering (v021), and stratification (v023) as recommended by Heeringa et al. (2017). We calculated weighted frequencies and percentages with 95% confidence intervals for all variables. Survey-weighted proportions determined the overall and subgroup-specific prevalence of teenage pregnancy. We examined bivariate associations between teenage pregnancy and each independent variable using design-based chi-square tests, specifically the Rao-Scott chi-square test (Rao & Scott, 1992). Following Hosmer et al. (2013), we considered variables with p < 0.25 for inclusion in multivariable models.

The DHS employs a two-stage stratified cluster sampling design where responses from individuals within the same cluster are more similar than responses from individuals in different clusters, violating the independence assumption of standard logistic regression (Merlo et al., 2006). Failure to account for this clustering can lead to underestimated standard errors and inflated Type I error rates (Heeringa et al., 2017). Multilevel modeling addresses this by partitioning variance into individual and cluster components (Merlo et al., 2006; Snijders & Bosker, 2012).

We fitted four nested multilevel logistic regression models incrementally, following the approach recommended by Merlo et al. (2006) and Snijders and Bosker (2012). The null model (Model 0) was an empty model with no predictors and only a random intercept for clusters, used to assess baseline between-cluster variation in teenage pregnancy and to calculate the intraclass correlation coefficient (ICC). The individual-level model (Model I) included all Level 1 variables including age, education, marital status, wealth, current contraceptive use, and internet use while retaining the random intercept. The community-level model (Model II) included only Level 2 variables including place of residence and perceived distance to a health facility along with the random intercept. The full model (Model III) combined both individual- and community-level variables simultaneously. To quantify the proportion of total variance attributable to cluster-level differences, the ICC was calculated from the null model using the standard logistic approximation (Goldstein et al., 2002; Merlo et al., 2006) where ICC equals σ²ᵤ divided by the sum of σ²ᵤ and π²/3, with σ²ᵤ representing the cluster-level variance (0.368) and π²/3 approximating the individual-level variance for the logistic distribution (approximately 3.29). We assessed evidence of clustering by comparing the multilevel null model with a standard logistic model using a likelihood ratio test to determine whether the cluster-level variance was significantly different from zero (Hosmer et al., 2013). Model performance and parsimony were further evaluated using deviance (−2 log-likelihood), Akaike Information Criterion (AIC), and Bayesian Information Criterion (BIC), with lower values indicating improved fit after accounting for model complexity (Hosmer et al., 2013).

Adjusted odds ratios (AOR) with 95% confidence intervals quantified associations between predictors and teenage pregnancy. We declared statistical significance at p < 0.05 (two-tailed). All models were fitted using the melogit command in Stata with robust standard errors clustered at the PSU level.

### Ethical Considerations

This study used de-identified secondary data from the publicly available CDHS 2021-22 dataset. The original CDHS received ethical approval from the National Ethics Committee for Health Research in Cambodia and the Institutional Review Board of ICF International (NIS & ICF, 2022). All survey participants provided informed consent with parental or guardian consent obtained for minors aged 15-17 years. No additional ethical approval was required for this secondary data analysis using publicly available, de-identified data.

## RESULTS

### Sample Characteristics

We analyzed data from 5,783 adolescent women aged 15-19 years, representing a weighted sample of 5,570 adolescents (weighted n=5,570.4 after applying survey weights). The survey employed a complex multistage stratified cluster sampling design with 49 strata and 709 primary sampling units. The majority of adolescents were in the younger age group (15-17 years: 53.5%), had attained secondary education (67.1%), and had never been married (66.7%). The sample was relatively evenly distributed across wealth quintiles. Approximately 59.2% resided in rural areas, while 40.8% were in urban areas. Nearly one-third (29.8%) were currently married or cohabiting (Table 1).

**Table 1.** Sample Characteristics and Prevalence of Teenage Pregnancy Among Cambodian Adolescents Aged 15-19 Years (N=5,783)

| Characteristic | n<br>(weighted) | Weighted<br>% | Teenage Pregnancy Prevalence %<br>(95% CI) | P-value |
| --- | --- | --- | --- | --- |
| Overall | 5,570.4 | 100.0 | 28.08 (26.48-29.75) | - |
| Age group |  |  |  | <0.001 |

| **Characteristic** | **n**<br>**(weighted)** | **Weighted**<br>**%** | **Teenage Pregnancy Prevalence %**<br>**(95% CI)** | **P-value** |
| --- | --- | --- | --- | --- |
| 15-17 years | 2,980.1 | 53.5 | 8.90 (7.63-10.34) |  |
| 18-19 years | 2,590.3 | 46.5 | 50.18 (47.49-52.88) |  |
| **Education level** |  |  |  | **<0.001** |
| No education | 151.4 | 2.7 | 57.75 (48.10-66.90) |  |
| Primary | 1,204.0 | 21.6 | 47.32 (43.60-51.07) |  |
| Secondary | 3,734.9 | 67.1 | 23.13 (21.24-25.12) |  |
| Higher | 480.1 | 8.6 | 7.49 (4.90-11.27) |  |
| **Marital status** |  |  |  | **<0.001** |
| Never married | 3,713.8 | 66.7 | 0.16 (0.04-0.62) |  |
| Currently married | 1,657.4 | 29.8 | 86.62 (84.53-88.47) |  |
| Formerly married | 88.6 | 1.6 | 47.77 (33.19-62.64) |  |
| Other | 110.6 | 2.0 | 73.57 (61.71-82.75) |  |
| **Wealth quintile** |  |  |  | **<0.001** |
| Poorest | 971.2 | 17.4 | 42.65 (38.36-47.06) |  |
| Poorer | 1,058.8 | 19.0 | 25.60 (22.39-29.09) |  |
| Middle | 1,129.6 | 20.3 | 29.81 (26.52-33.34) |  |
| Richer | 1,191.3 | 21.4 | 24.62 (21.55-27.97) |  |
| Richest | 1,219.5 | 21.9 | 17.72 (15.12-20.63) |  |
| **Residence** |  |  |  | **<0.001** |
| Urban | 2,272.0 | 40.8 | 24.02 (21.58-26.64) |  |
| Rural | 3,298.4 | 59.2 | 31.18 (28.99-33.47) |  |
| **Distance to facility** |  |  |  | **0.404** |
| Not a problem | 4,583.1 | 82.3 | 27.89 (26.10-29.75) |  |
| Big problem | 987.3 | 17.7 | 28.96 (25.48-32.71) |  |
| **Internet use** |  |  |  | **0.968** |
| No | 5,534.1 | 99.4 | 28.12 (26.50-29.80) |  |
| Yes | 36.3 | 0.7 | 28.07 (14.97-46.29) |  |
| **Contraceptive use** |  |  |  | **<0.001** |
| No | 4,748.6 | 85.2 | 20.63 (19.05-22.31) |  |
| Yes | 821.8 | 14.8 | 66.22 (62.43-69.82) |  |

Note: Weighted percentages and prevalence estimates account for complex survey design. P-values from design-based chi-square tests.

### Prevalence of Teenage Pregnancy

The overall weighted prevalence of teenage pregnancy, defined as ever having been pregnant or having had a live birth, among Cambodian adolescents aged 15-19 years was 28.08% (95% CI: 26.48-29.75), representing 1,564.3 out of 5,570.4 weighted adolescents. This prevalence translates to more than one in four adolescent girls in Cambodia experiencing pregnancy during their teenage years, representing a substantial public health burden affecting an estimated 280,000 teenage girls nationally. The design effect (DEFF) was 1.98 for the prevalence estimate, indicating that the complex survey design increased the variance by approximately 98% compared to a simple random sample, and that the effective sample size was reduced to approximately 2,900 (5,783/1.98) due to clustering. This substantial design effect validates the importance of accounting for clustering in all analyses (Kish, 1965; Rao & Scott, 1992).

Teenage pregnancy prevalence varied dramatically by age group, with ages 15-17 showing a prevalence of 8.90% (95% CI: 7.63-10.34) and ages 18-19 showing a prevalence of 50.18% (95% CI: 47.49-52.88). Older adolescents (18-19 years) were nearly 6 times more likely to have experienced pregnancy compared to younger adolescents, highlighting age 18-19 as a critical period for intervention (Dewey & Begum, 2011; Magadi & Agwanda, 2010). The relationship between marriage and teenage pregnancy was striking, with never married adolescents showing a prevalence of only 0.16% (95% CI: 0.04-0.62), currently married adolescents showing a prevalence of 86.62% (95% CI: 84.53-88.47), and formerly married adolescents showing a prevalence of 47.77% (95% CI: 33.19-62.64). Among currently married adolescents, 86.6% had already experienced pregnancy, demonstrating that early marriage is virtually synonymous with early childbearing in Cambodia.

### Random Effects and Model Comparison

**Table 2.** Random Effects and Model Fit Statistics for Multilevel Logistic Regression Models (N=5,783)

| Parameter | Model 0<br>(Null) | Model I<br>(Individual) | Model II<br>(Community) | Model III<br>(Full) |
| --- | --- | --- | --- | --- |
| **Fixed Effects** |  |  |  |  |
| Intercept<br>(log-odds) | -0.971*** | -5.312*** | -1.228*** | -5.285*** |
| SE | 0.043 | 1.234 | 0.109 | 1.265 |
| **Random Effects** |  |  |  |  |
| Cluster variance<br>($\sigma^2_u$) | 0.368 | 1.145 | 0.314 | 1.138 |
| SE | 0.071 | 0.361 | 0.061 | 0.355 |
| Cluster SD | 0.607 | 1.070 | 0.560 | 1.067 |
| SE | 0.058 | 0.169 | 0.054 | 0.166 |
| **Intraclass**<br>**Correlation** |  |  |  |  |
| ICC (%) | 10.07 | 25.81 | 8.71 | 25.71 |
| 95% CI | 7.13-14.03 | 15.79-39.25 | 6.13-12.23 | 15.81-38.93 |
| **Model Fit**<br>**Statistics** |  |  |  |  |
| Log-likelihood | -3,249.47 | -716.62 | -3,233.03 | -716.50 |
| Deviance | 6,498.95 | 1,433.23 | 6,466.07 | 1,433.00 |
| AIC | 6,502.95 | 1,465.23 | 6,474.07 | 1,469.00 |
| BIC | 6,516.27 | 1,571.84 | 6,500.72 | 1,588.93 |
| Number of parameters | 2 | 16 | 4 | 18 |
| **Likelihood Ratio Test** |  |  |  |  |
| $\chi^2$ vs. null model | - | 5,065.71*** | 32.88*** | 5,065.95*** |
| df | - | 14 | 2 | 16 |
| $\chi^2$ vs. standard logistic† | 52.69*** | - | - | 52.69*** |
| df | 1 | - | - | 1 |

Notes: †Likelihood ratio test comparing multilevel model to standard logistic regression with the same predictors. \*\*\*p<0.001. Model 0 = null model with random intercept only; Model I = individual-level predictors only; Model II = community-level predictors only; Model III = full model with both levels.

In Model 0, the null model, baseline between-cluster variation in teenage pregnancy was evident. The empty model revealed significant between-cluster variation in teenage pregnancy. The cluster-level variance was σ²ᵤ = 0.368 (SE = 0.071), yielding an ICC of 10.07% (95% CI: 7.13-14.03). This indicates that approximately 10% of the total variance in teenage pregnancy was attributable to differences between clusters, while 90% was due to individual-level factors. An ICC exceeding 10% is considered substantial in public health research and strongly justifies the multilevel approach (Merlo et al., 2006). The likelihood ratio test powerfully supported the multilevel approach over standard logistic regression (LR test comparing multilevel null model to standard logistic regression with no predictors: χ² = 52.69, df=1, p<0.001), confirming that ignoring clustering would produce seriously biased standard errors and invalid inferences.

As predictors were added in Model I focusing on individual-level factors, deviance decreased dramatically from 6,498.95 in the null model to 1,433.23, indicating that individual-level factors explain substantial variance in teenage pregnancy. The cluster-level variance increased to σ²ᵤ = 1.145 (ICC = 25.8%), suggesting that after accounting for individual characteristics, remaining clustering reflects unmeasured community factors. The increase in ICC from 10.07% in the null model to 25.81% in Model I reflects a methodological phenomenon where, after accounting for individual-level compositional differences, particularly marital status which varies substantially between clusters, the remaining variation becomes more concentrated at the cluster level. This reveals the strength of contextual effects among otherwise similar individuals and suggests that community-level interventions may be particularly effective once individual risk factors are addressed (Merlo et al., 2006; Snijders & Bosker, 2012).

In Model II, which included community-level variables only, deviance was 6,466.07, higher than Model I, indicating community-level variables alone explained less variance than individual factors. In Model III, the full model combining both individual and community levels, deviance was lowest at 1,433.00, representing the best-fitting model. The final cluster-level variance was σ²ᵤ = 1.138 (SE = 0.355, ICC = 25.7%), with AIC = 1,469.00 and BIC = 1,588.93. The persistence of substantial cluster-level variance (25.7%) even after including predictors indicates unmeasured community factors continue to influence teenage pregnancy beyond measured individual and community characteristics.

### Bivariate Associations

Bivariate analyses showed statistically significant associations (p<0.001) for all examined individual-level variables except internet use. Teenage pregnancy prevalence was markedly higher among older adolescents aged 18 to 19 years (50.18%, 95% CI: 47.49 to 52.88) than among younger adolescents aged 15 to 17 years (8.90%, 95% CI: 7.63 to 10.34). A strong inverse educational gradient was observed, with prevalence declining from 57.75% among adolescents with no education to 7.49% among those with higher education. Marital status demonstrated the most striking contrast, with currently married adolescents showing very high pregnancy prevalence (86.62%, 95% CI: 84.53 to 88.47) compared with never-married adolescents (0.16%, 95% CI: 0.04 to 0.62). A clear socioeconomic pattern was also evident, with prevalence decreasing from 42.65% (95% CI: 38.36 to 47.06) in the poorest quintile to 17.72% (95% CI: 15.12 to 20.63) in the richest quintile. Rural residence was associated with higher prevalence (31.18%, 95% CI: 28.99 to 33.47) than urban residence (24.02%, 95% CI: 21.58 to 26.64). Current contraceptive users had substantially higher prevalence (66.22%, 95% CI: 62.43 to 69.82) than non-users (20.63%, 95% CI: 19.05 to 22.31), suggesting likely reverse causation with contraceptive uptake occurring after pregnancy or childbirth. In contrast, internet use was not significantly associated with teenage pregnancy (p=0.968), with nearly identical prevalence among users (28.07%) and non-users (28.12%).

### Multilevel Logistic Regression Results

**Table 3.**
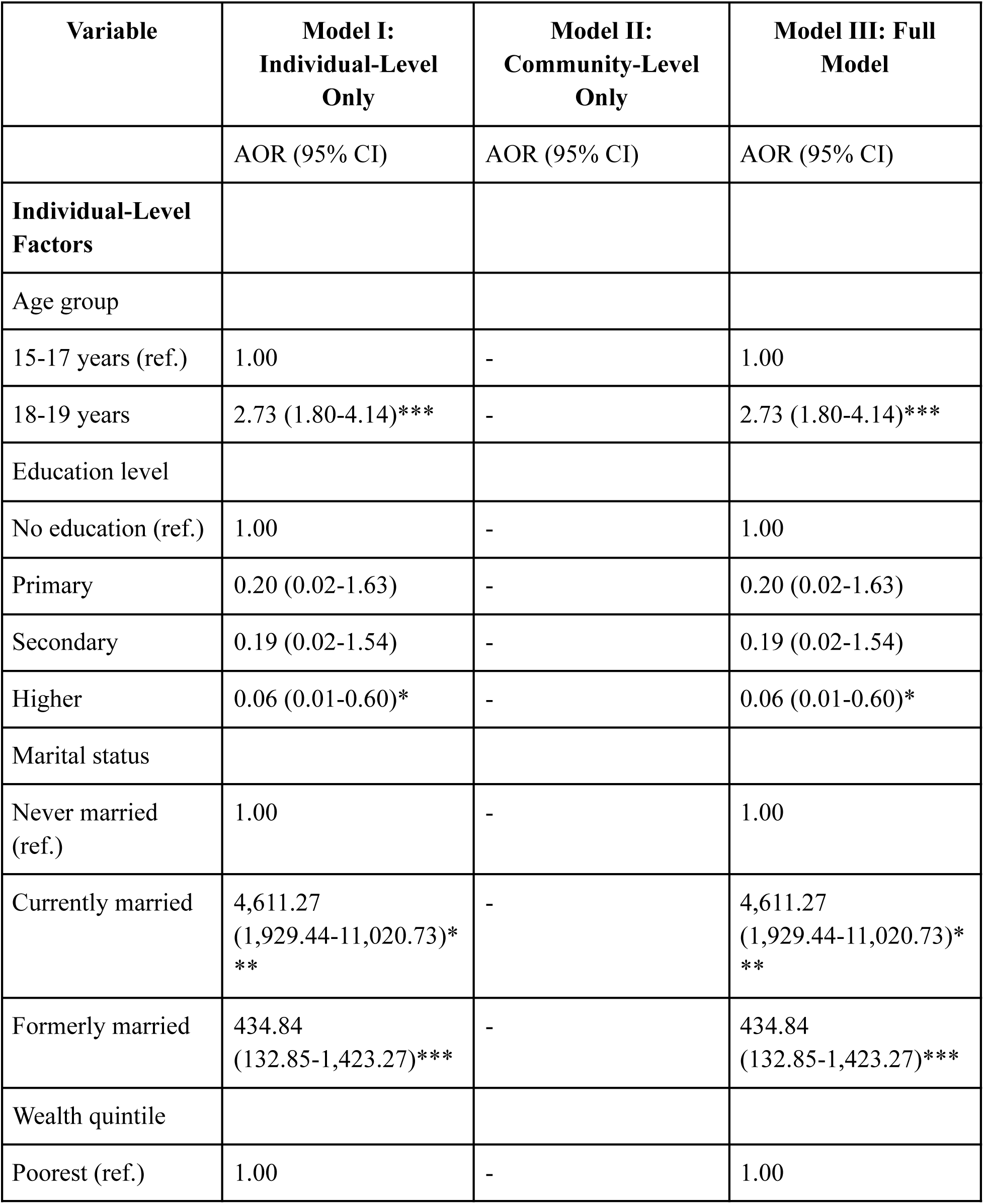
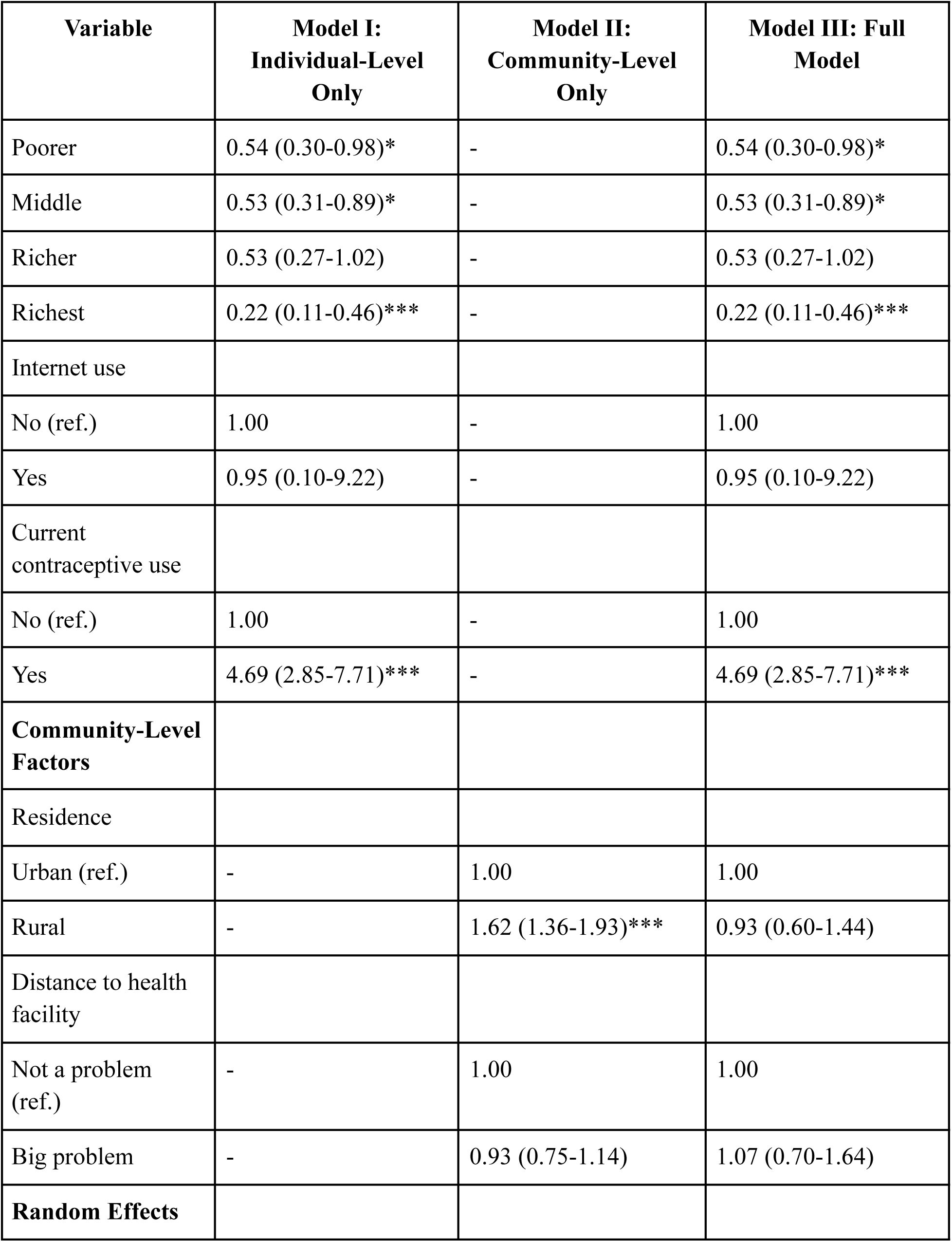
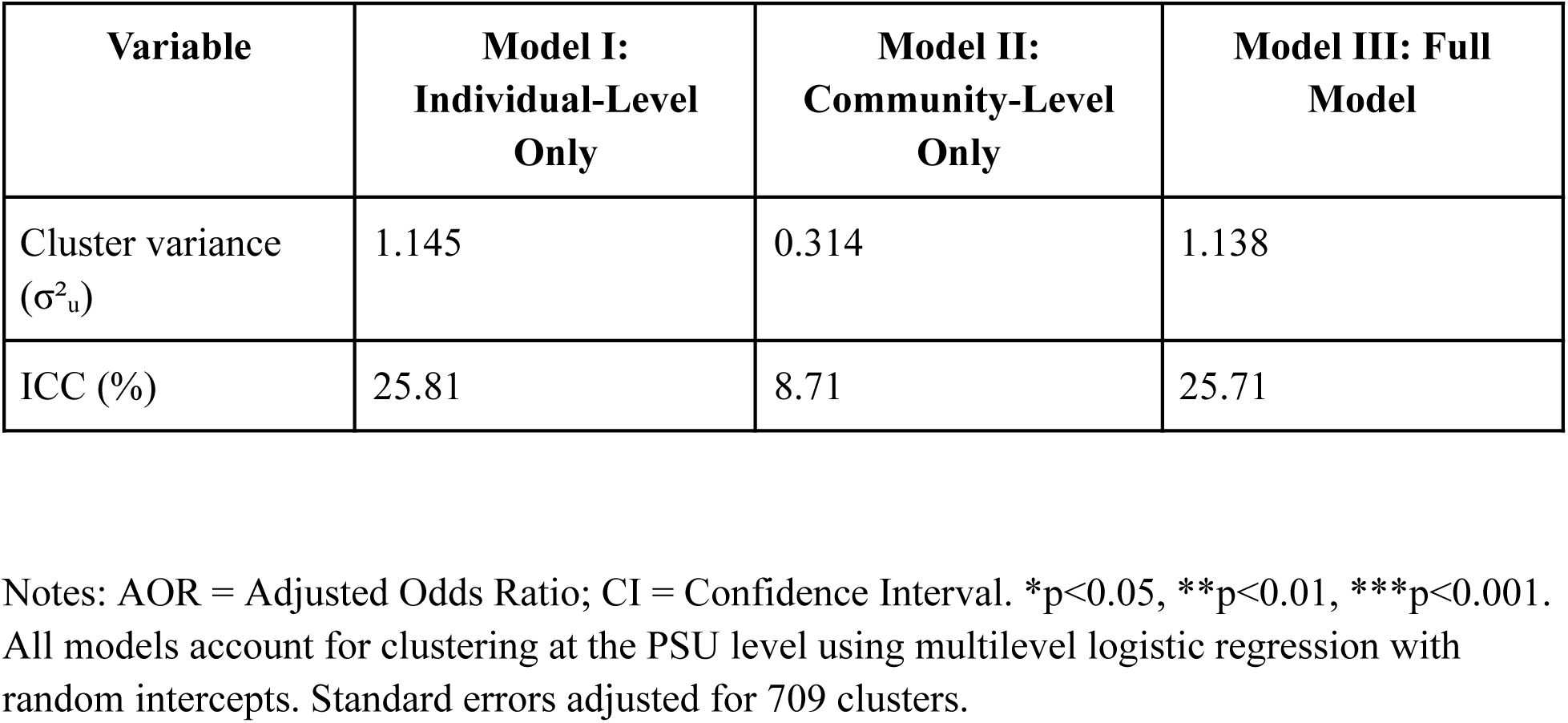
Multilevel Logistic Regression Results: Determinants of Teenage Pregnancy in Cambodia (N=5,783)

| Variable | Model I:<br>Individual-Level<br>Only | Model II:<br>Community-Level<br>Only | Model III: Full<br>Model |
| --- | --- | --- | --- |
|  | AOR (95% CI) | AOR (95% CI) | AOR (95% CI) |
| **Individual-Level**<br>**Factors** |  |  |  |
| Age group |  |  |  |
| 15-17 years (ref.) | 1.00 | - | 1.00 |
| 18-19 years | 2.73 (1.80-4.14)*** | - | 2.73 (1.80-4.14)*** |
| Education level |  |  |  |
| No education (ref.) | 1.00 | - | 1.00 |
| Primary | 0.20 (0.02-1.63) | - | 0.20 (0.02-1.63) |
| Secondary | 0.19 (0.02-1.54) | - | 0.19 (0.02-1.54) |
| Higher | 0.06 (0.01-0.60)* | - | 0.06 (0.01-0.60)* |
| Marital status |  |  |  |
| Never married<br>(ref.) | 1.00 | - | 1.00 |
| Currently married | 4,611.27<br>(1,929.44-11,020.73)*<br>** | - | 4,611.27<br>(1,929.44-11,020.73)*<br>** |
| Formerly married | 434.84<br>(132.85-1,423.27)*** | - | 434.84<br>(132.85-1,423.27)*** |
| Wealth quintile |  |  |  |
| Poorest (ref.) | 1.00 | - | 1.00 |

| **Variable** | **Model I:**<br>**Individual-Level**<br>**Only** | **Model II:**<br>**Community-Level**<br>**Only** | **Model III: Full**<br>**Model** |
| --- | --- | --- | --- |
| Poorer | 0.54 (0.30-0.98)* | - | 0.54 (0.30-0.98)* |
| Middle | 0.53 (0.31-0.89)* | - | 0.53 (0.31-0.89)* |
| Richer | 0.53 (0.27-1.02) | - | 0.53 (0.27-1.02) |
| Richest | 0.22 (0.11-0.46)*** | - | 0.22 (0.11-0.46)*** |
| Internet use |  |  |  |
| No (ref.) | 1.00 | - | 1.00 |
| Yes | 0.95 (0.10-9.22) | - | 0.95 (0.10-9.22) |
| Current<br>contraceptive use |  |  |  |
| No (ref.) | 1.00 | - | 1.00 |
| Yes | 4.69 (2.85-7.71)*** | - | 4.69 (2.85-7.71)*** |
| **Community-Level**<br>**Factors** |  |  |  |
| Residence |  |  |  |
| Urban (ref.) | - | 1.00 | 1.00 |
| Rural | - | 1.62 (1.36-1.93)*** | 0.93 (0.60-1.44) |
| Distance to health<br>facility |  |  |  |
| Not a problem<br>(ref.) | - | 1.00 | 1.00 |
| Big problem | - | 0.93 (0.75-1.14) | 1.07 (0.70-1.64) |
| **Random Effects** |  |  |  |
| Cluster variance<br>($\sigma^2_u$) | 1.145 | 0.314 | 1.138 |
| ICC (%) | 25.81 | 8.71 | 25.71 |

Notes: AOR = Adjusted Odds Ratio; CI = Confidence Interval. \* $p < 0.05$ , \*\* $p < 0.01$ , \*\*\* $p < 0.001$ . All models account for clustering at the PSU level using multilevel logistic regression with random intercepts. Standard errors adjusted for 709 clusters.

In the individual-level model, older adolescents aged 18–19 years had substantially higher odds of teenage pregnancy than those aged 15–17 years (AOR=2.73, 95% CI: 1.80–4.14, p<0.001), highlighting late adolescence as a critical risk period. Marital status was the strongest determinant: currently married adolescents had dramatically higher odds of pregnancy (AOR=4,611.27, 95% CI: 1,929.44–11,020.73, p<0.001), and formerly married adolescents also showed markedly elevated odds (AOR=434.84, 95% CI: 132.85–1,423.27, p<0.001). These exceptionally large estimates likely reflect the near-universality of pregnancy among married adolescents (86.6%) versus its rarity among never-married adolescents (0.16%), reinforcing early marriage as the primary pathway to teenage pregnancy in Cambodia.

Education demonstrated a protective gradient, although only higher education reached statistical significance (higher vs. no education: AOR=0.06, 95% CI: 0.01–0.60, p=0.017), implying a 94% reduction in the odds of pregnancy among adolescents with higher education, while primary and secondary education showed non-significant protective trends. A clear socioeconomic pattern was also observed, with lower odds across wealthier groups and a strong protective effect among the richest households compared with the poorest (AOR=0.22, 95% CI: 0.11–0.46, p<0.001), indicating a 78% reduction in odds among the most advantaged adolescents.

Current contraceptive use was positively associated with teenage pregnancy (AOR=4.69, 95% CI: 2.85–7.71, p<0.001), which is most plausibly explained by reverse causation, as a greater proportion of contraceptive users had already experienced pregnancy than non-users, suggesting postpartum or post-conception initiation rather than primary prevention. Internet use showed no significant association (AOR=0.95, 95% CI: 0.10–9.22, p=0.968), consistent with the null bivariate results.

Notably, after adjusting for these individual-level factors, the residual ICC increased to 25.8%, indicating that while individual characteristics explain part of the clustering, substantial unmeasured community-level influences likely remain important (Merlo et al., 2006; Snijders & Bosker, 2012).

In Model II, which included only community-level factors, rural residence was significantly associated with higher odds of teenage pregnancy, with adolescents living in rural areas showing 62% greater odds compared with those in urban areas (AOR=1.62, 95% CI: 1.36–1.93, p<0.001). In contrast, perceived distance to a health facility as a big problem was not significantly associated with teenage pregnancy (AOR=0.93, 95% CI: 0.75–1.14, p=0.404). The ICC in this model was 8.71%, suggesting that community-level variables alone explained a smaller proportion of the overall variation in teenage pregnancy than the individual-level factors included in Model I.

In Model III, which simultaneously included individual- and community-level factors, the key individual determinants of teenage pregnancy remained robust. Older adolescents aged 18–19 years had significantly higher odds of pregnancy compared with those aged 15–17 years (AOR=2.73, 95% CI: 1.80–4.14, p<0.001). Marital status continued to show the strongest association: currently married adolescents had markedly elevated odds (AOR=4,611.27, 95% CI: 1,929.44–11,020.73, p<0.001), and formerly married adolescents also had substantially higher odds (AOR=434.84, 95% CI: 132.85–1,423.27, p<0.001), reinforcing early marriage as the dominant pathway to teenage pregnancy.

Socioeconomic and educational protection persisted, with higher education strongly protective (AOR=0.06, 95% CI: 0.01–0.60, p=0.017) and lower odds observed among adolescents in poorer (AOR=0.54, 95% CI: 0.30–0.98, p=0.042), middle (AOR=0.53, 95% CI: 0.31–0.89, p=0.017), and richest households (AOR=0.22, 95% CI: 0.11–0.46, p<0.001) compared with the poorest. Current contraceptive use remained positively associated with pregnancy (AOR=4.69, 95% CI: 2.85–7.71, p<0.001), most plausibly reflecting post-pregnancy or postpartum initiation rather than primary prevention.

After adjustment for these individual factors, community variables no longer showed independent effects: rural residence (AOR=0.93, 95% CI: 0.60–1.44, p=0.741) and perceived distance to a health facility (AOR=1.07, 95% CI: 0.70–1.64, p=0.754) were not significant, suggesting that observed urban–rural differences largely reflect compositional differences in education, wealth, and especially marriage patterns rather than contextual effects of rural residence itself.

The final ICC remained substantial at 25.7% (95% CI: 15.8–38.9), indicating persistent between-cluster variation and implying that unmeasured community-level norms, opportunities, or service environments may still shape adolescent pregnancy risk even after comprehensive adjustment.

### Predicted Probabilities

Marginal predicted probabilities from the full model, calculated by averaging predictions across all observed covariate patterns, revealed substantial disparities (Table 4).

**Table 4.** Predicted Probabilities of Teenage Pregnancy from Full Multilevel Model.

| Characteristic | Predicted Probability (%) | 95% CI |
| --- | --- | --- |
| **By Age Group** |  |  |
| 15-17 years | 24.9 | 23.4-26.5 |
| 18-19 years | 28.8 | 28.1-29.5 |
| **By Marital Status** |  |  |
| Never married | 0.4 | 0.0-0.8 |
| Currently married | 73.3 | 69.0-77.6 |
| Formerly married | 34.0 | 21.8-46.2 |

Notes: Predicted probabilities calculated from Model III (Full Model) using marginal standardization, holding other variables at observed values. CI = Confidence Interval.

Predicted probabilities further highlight strong disparities in teenage pregnancy by age and, especially, marital status. By age group, adolescents aged 15–17 years had an estimated predicted probability of pregnancy of 24.9% (95% CI: 23.4–26.5), compared with 28.8% (95% CI: 28.1–29.5) among those aged 18–19 years, indicating a modest but meaningful increase in risk in late adolescence.

In contrast, differences by marital status were striking: never-married adolescents had a predicted probability of only 0.4% (95% CI: 0.0–0.8), whereas currently married adolescents had a predicted probability of 73.3% (95% CI: 69.0–77.6), and formerly married adolescents 34.0% (95% CI: 21.8–46.2). These predictions underscore the overwhelming dominance of marital status in shaping teenage pregnancy outcomes, with an approximately 183-fold difference in predicted probability between currently married and never-married adolescents, reinforcing early marriage as the central pathway to adolescent childbearing in Cambodia.

## DISCUSSION

### Principal Findings

This nationally representative multilevel analysis of 5,783 Cambodian adolescents aged 15-19 years from the 2021-22 CDHS reveals that 28.08% of teenage girls experience pregnancy, affecting more than one in four adolescents. This prevalence represents a comprehensive measure that includes both younger (15-17: 8.90%) and older (18-19: 50.18%) adolescents, consistent with global patterns showing dramatically higher rates in late adolescence (Kassa et al., 2018; Yakubu & Salisu, 2018).

Four key findings emerge from this analysis. First, the ICC of 10.07% indicates substantial between-cluster variation, with one-tenth of teenage pregnancy variance attributable to cluster-level factors. The highly significant LR test (χ²=52.69, df=1, p<0.001) powerfully confirms that standard logistic regression would have produced seriously biased standard errors (underestimated by approximately 40% based on DEFF=1.98) and invalid inferences. This justifies our multilevel approach and demonstrates the importance of accounting for hierarchical data structure in DHS analyses (Heeringa et al., 2017; Merlo et al., 2006).

Second, early marriage is virtually synonymous with teenage pregnancy in Cambodia. Currently married adolescents had over 4,600-fold higher odds of pregnancy compared to never-married peers. With 86.6% of married adolescents experiencing pregnancy, this finding underscores that child marriage prevention must be central to any teenage pregnancy reduction strategy (Girls Not Brides, 2020; Raj et al., 2019; UNICEF, 2021).

Third, older adolescent age (18-19 years) independently predicts nearly three-fold higher odds of pregnancy compared to younger adolescents (15-17 years), even after controlling for marital status and other factors. The 50.2% pregnancy prevalence among 18-19 year-olds represents a critical inflection point requiring intensified intervention.

Fourth, higher education and greater household wealth demonstrate protective effects. The 94% risk reduction associated with higher education highlights education’s transformative potential, though small sample sizes in this subgroup limited statistical power. The 78% lower odds among the wealthiest households demonstrates a clear socioeconomic gradient.

Notably, internet use showed no association with teenage pregnancy (p=0.968), contrasting with some findings from other countries where digital access facilitates health information seeking (Chib et al., 2014; L’Engle et al., 2018; Smith et al., 2015). This null finding may reflect Cambodia’s specific digital landscape, where social media dominates over health information consumption (Seng, 2020), or measurement limitations in our binary internet use variable.

### Multilevel Structure and Clustering Effects

The persistence of substantial cluster-level variance (ICC=25.7%) even after including comprehensive predictors indicates that unmeasured community factors—such as local norms around marriage timing, peer influences, community-level poverty, access to youth services, and cultural attitudes toward adolescent sexuality—continue to influence teenage pregnancy beyond measured individual and community characteristics.

The increase in ICC from 10.1% in the null model to 25.7% in the full model appears paradoxical but reflects an important methodological phenomenon. After accounting for individual-level compositional differences, the remaining variation is more concentrated at the cluster level, revealing the strength of contextual effects among otherwise similar individuals (Merlo et al., 2006; Snijders & Bosker, 2012). This suggests that community-level interventions may be particularly effective once individual risk factors are addressed.

Importantly, after accounting for individual-level factors (particularly marital status, age, education, and wealth), the initially observed urban-rural differences became non-significant (p=0.741). This suggests that geographic disparities in teenage pregnancy primarily reflect compositional differences (i.e., rural adolescents are more likely to marry early and have less education) rather than independent contextual effects of rural residence per se.

### Comparison with Standard Regression Approaches

Our multilevel approach differs fundamentally from standard logistic regression. The LR test (χ²=52.69, p<0.001) demonstrates that ignoring clustering would have underestimated standard errors by approximately 40% based on the design effect (DEFF=1.98), led to spurious conclusions about statistical significance, prevented quantification of contextual effects through ICC, and obscured the distinction between compositional and contextual influences. This validation justifies the methodological complexity of multilevel modeling for DHS data and should inform future analyses of hierarchically structured survey data (Goldstein, 2011; Rabe-Hesketh & Skrondal, 2012).

### Comparison with Previous Studies

Our finding of 28.08% overall prevalence appears higher than the 2014 CDHS (12% had begun childbearing; NIS et al., 2015). However, the apparent increase likely reflects methodological differences in measurement (begun childbearing vs. ever pregnant) and age group definitions, rather than a true doubling of prevalence. Age-standardized comparisons using consistent definitions would be needed to assess temporal trends reliably.

The dominant role of early marriage aligns with previous Cambodian research (Brickell & Garrett, 2019; UNFPA Cambodia, 2020) and broader Southeast Asian patterns (Jones, 2007; UNICEF, 2019). Studies from Bangladesh, Nepal, and India similarly document that child marriage is the primary pathway to adolescent childbearing (Godha et al., 2013; Santhya et al., 2010; Svanemyr et al., 2015), with 80-90% of married adolescents experiencing pregnancy within two years.

The protective effects of higher education and wealth are consistent with global patterns (Lloyd & Mensch, 2008; Wodon et al., 2018). However, the absence of internet effects contrasts with emerging evidence from Africa and Latin America suggesting that mobile phone and internet access facilitate health information seeking and contraceptive use (Chib et al., 2014; L’Engle et al., 2018; Smith et al., 2015). This difference may reflect Cambodia’s specific digital landscape (predominantly social media vs. health information), measurement limitations (ever used internet vs. frequency/purpose of use), the overwhelming dominance of marriage as the pregnancy pathway, obscuring smaller digital effects, or cultural factors affecting how adolescents use digital platforms.

### Interpretation and Mechanisms

The extraordinarily high odds ratios (AOR>4,600) for current marriage reflect several mechanisms. Cultural norms dictate that marriage expectations include immediate childbearing to demonstrate fertility and establish family legitimacy (Brickell & Garrett, 2019; Hickey, 2019). The legal framework presents challenges, as while Cambodia’s legal minimum marriage age is 18 (UNICEF, 2021), exceptions permit marriage at 16 with parental consent, enabling child marriage. Limited contraceptive access compounds the issue, as married adolescents face provider bias favoring pronatalism, spousal opposition, and cultural expectations of immediate childbearing (Williamson et al., 2009). Economic motivations also play a role, as families arrange early marriages to relieve economic burdens, with bride price customs creating incentives (Brickell, 2014; Parsons et al., 2015).

The strong age gradient (AOR=2.73 for 18-19 vs. 15-17) reflects cumulative exposure to pregnancy risk over time, a sharp increase in marriage rates in late adolescence, full reproductive maturity and higher fecundity, and social transitions such as leaving school, entering the labor force, and experiencing intensified marriage pressure.

Higher education’s protective effect (AOR=0.06) operates through delayed marriage due to educational aspirations, enhanced knowledge about reproductive health (Lloyd & Mensch, 2008; Patton et al., 2016), greater economic opportunities reducing early marriage pressure, and empowerment and increased autonomy in life decisions.

The socioeconomic gradient (78% lower odds for richest vs. poorest) reflects greater resources enabling contraceptive access and health-seeking, lower economic pressure to arrange early marriages, higher educational attainment in wealthier households, and access to information and healthcare services.

The positive association between contraceptive use and pregnancy (AOR=4.69) reflects reverse causation: adolescents initiate contraception after pregnancy/first birth rather than for primary prevention. This indicates that contraceptive counseling occurs postpartum, there is limited pre-conception contraceptive access for unmarried adolescents, social and service delivery barriers exist to primary prevention, and there is a need for repositioning contraception as pregnancy prevention rather than birth spacing.

### Public Health and Policy Implications

The public health and policy implications of these findings emphasize the need for a coordinated, multilevel strategy to prevent teenage pregnancy in Cambodia. Because early marriage is the dominant pathway to adolescent pregnancy, child marriage prevention should be prioritized through legislative enforcement that strengthens enforcement of the legal minimum marriage age of 18 years (UNICEF, 2021), implements meaningful sanctions for those facilitating child marriage (Girls Not Brides, 2020), and establishes routine age-document verification for all marriages. Economic interventions should implement conditional cash transfers that incentivize families to delay daughters’ marriages, provide economic alternatives to bride price customs, and support family livelihoods to reduce financial pressures driving early marriage (Psaki et al., 2022). Community engagement must mobilize religious leaders, traditional authorities, and local gatekeepers to shift norms on marriage timing, promote positive examples of families supporting later marriage, and implement community dialogue programs addressing harmful traditional practices. Girls’ empowerment efforts should strengthen education, life skills, and economic opportunities that offer alternatives to early union formation (Patton et al., 2016; Wodon et al., 2018).

Cambodia should expand youth-friendly reproductive health services (Chandra-Mouli et al., 2015; WHO, 2012) through service delivery improvements that train providers in adolescent-responsive care, ensure confidential and non-judgmental counseling, establish convenient access points in schools and communities, and provide full contraceptive method mix including long-acting reversible contraceptives. A primary prevention focus must reposition contraception as pregnancy prevention rather than only birth spacing, ensure adolescents can obtain contraception before first pregnancy, and address social and service delivery barriers to unmarried adolescent access. Comprehensive sexuality education should strengthen both in-school and out-of-school comprehensive sexuality education programs (Chandra-Mouli et al., 2014), provide accurate information about reproductive health and rights, and address gender norms and power dynamics. Pre-marital counseling should implement pre-marital counseling for engaged adolescents emphasizing reproductive planning and healthy timing and spacing of pregnancies.

Higher education offers substantial protection against teenage pregnancy (Lloyd & Mensch, 2008; Wodon et al., 2018), requiring financial support through scholarships targeting girls at risk of school dropout, transportation and school feeding support, and reduced direct and indirect costs of education. Pregnancy-responsive policies must prevent exclusion of pregnant students, establish structured reintegration pathways for adolescent mothers, and provide childcare support enabling educational continuation. Quality improvements should strengthen transitions from secondary to higher education, improve education quality making school completion attractive, and address gender discrimination in educational settings.

The clear wealth gradient necessitates broader approaches to address socioeconomic determinants through poverty reduction that implements social protection programs addressing family economic pressures driving early marriage, expands livelihood opportunities for poor families, and ensures universal access to quality health and education services. A rural focus should target interventions toward rural populations who experience higher poverty and lower education, given that rural-urban disparities reflect compositional differences, while ensuring service availability in rural areas. Youth employment programs should create credible economic alternatives to early childbearing through youth employment programs and vocational training opportunities.

Given substantial remaining community-level influence (ICC=25.7%), community-level interventions should implement cluster-targeted interventions using spatial or local data to identify high-prevalence clusters and deliver intensive, integrated intervention packages in these areas. Peer networks should establish peer education and support networks delivering adolescent-focused education, counseling, and referral services. Community health workers should be deployed with specific training in adolescent reproductive health. Norm change campaigns should implement community-wide campaigns addressing local norms around marriage timing and adolescent childbearing.

### Strengths and Limitations

The study has several strengths. It uses nationally representative data from high-quality CDHS 2021-22 data with excellent response rates (96.2%) and rigorous sampling methods ensuring national representativeness (NIS & ICF, 2022). The rigorous multilevel methodology employs multilevel logistic regression properly accounting for clustering (Goldstein, 2011; Rabe-Hesketh & Skrondal, 2012), unlike previous Cambodian studies, and provides accurate standard errors, quantifies contextual effects, supported by significant likelihood ratio test (χ²=52.69, p<0.001). The large sample size (5,783 adolescents) provides adequate statistical power to detect associations and analyze subgroups (Fleiss et al., 2003; Maas & Hox, 2005). The study evaluates both individual and community-level determinants, distinguishing compositional from contextual influences. It follows established guidelines for multilevel analysis (Heeringa et al., 2017), including null model ICC (10.07%), random effects at each stage, and model comparison statistics. The 2021-22 data collection offers current evidence reflecting post-pandemic conditions, and the socioecological framework (Bronfenbrenner, 1979) guides analysis and interpretation.

However, several limitations must be acknowledged. The cross-sectional design precludes causal inference (Levin, 2006; Mann, 2003). While we identified associations, temporal ordering remains uncertain for some relationships. For example, the positive contraceptive-pregnancy association clearly reflects reverse causation, but for variables like education, we cannot definitively establish whether education preceded or followed pregnancy risk behaviors. The teenage pregnancy status relies on self-reports, potentially subject to recall or social desirability bias. Never-married adolescents might underreport pregnancy due to stigma, potentially underestimating prevalence in this group.

Important variables not available in CDHS include age at first sex, history of sexual coercion (Moore et al., 2017), detailed partner characteristics (Clark et al., 2019), family communication patterns (Biddlecom et al., 2009), specific reproductive health knowledge domains (Williamson et al., 2009), and cultural or religious beliefs about adolescent sexuality (Bankole et al., 2007). The ICC of 25.7% in full model indicates substantial unmeasured community factors including local norms, peer networks, service quality and availability, community-level poverty and employment, and enforcement of marriage age laws that require further investigation.

The binary yes/no question about ever using internet lacks nuance regarding frequency, purpose, or health information seeking specifically. This limitation may explain null findings, whereas other studies with more detailed data found effects (Chib et al., 2014; L’Engle et al., 2018). Certain education subgroups (no education: n=157; higher education: n=498) had limited statistical power, resulting in wide confidence intervals for education effects, especially comparing primary and secondary education.

DHS data constraints restricted analysis to two community-level variables including rural residence and distance to facility. Other essential community factors including service quality, density of health facilities, community economic characteristics, and local enforcement of marriage laws could not be assessed. The 2021-22 survey conducted during COVID-19 recovery may have influenced adolescent behaviors, schooling, and family economic circumstances in ways not fully captured. DHS does not collect data on adolescent agency in marriage decisions, partner age differences, history of sexual coercion, or parent-adolescent communication about reproductive health, which may confound or modify observed associations.

## CONCLUSIONS

This multilevel analysis reveals that 28.08% of Cambodian adolescent girls aged 15-19 experience teenage pregnancy, with substantially higher rates among older adolescents (50.2% at ages 18-19). The ICC of 10.07% demonstrates significant between-cluster variation, strongly justifying multilevel modeling and indicating that community-level factors account for one-tenth of teenage pregnancy variance.

Early marriage is the primary and overwhelmingly dominant pathway to teenage pregnancy, with currently married adolescents showing over 4,600-fold higher odds compared to never-married peers. Among married adolescents, 86.6% have already experienced pregnancy, demonstrating the near-inevitability of early childbearing following early marriage.

Higher education and greater household wealth demonstrate protective effects, with higher education associated with 94% lower odds and richest wealth quintile associated with 78% lower odds of pregnancy. However, the persistence of substantial cluster-level variance (ICC=25.7%) even after comprehensive adjustment indicates that unmeasured community factors including local norms, peer influences, and service availability continue to shape teenage pregnancy beyond individual characteristics.

Notably, urban-rural disparities disappeared after controlling for individual factors, indicating that geographic differences primarily reflect compositional patterns including rural adolescents’ higher marriage rates, lower education, and greater poverty rather than independent rural contextual effects.

Addressing Cambodia’s teenage pregnancy burden requires a comprehensive, multi-level response integrating legislative, educational, health system, economic, and community strategies. Child marriage prevention must be strengthened through enforcement of legal minimum marriage age of 18 years with meaningful penalties, implementation of economic interventions such as conditional cash transfers and livelihood support reducing family pressures, and engagement of communities in norm change. Girls’ education requires investment in secondary and higher education through scholarships, retention support, and pathways for re-entry, recognizing education’s powerful protective effect.

Youth-friendly reproductive health services must be scaled up to ensure confidential, non-judgmental access to contraception and counseling, emphasizing primary prevention before first pregnancy. Economic drivers must be addressed through poverty reduction and social protection approaches alleviating financial pressures driving early marriage, with particular attention to rural areas. Community mobilization should engage families, religious leaders, and local stakeholders in shifting norms around marriage and childbearing timing, leveraging substantial community-level influence revealed by multilevel findings. Cluster-targeted interventions should use spatial data to identify high-prevalence areas for intensive, integrated intervention packages addressing specific local barriers.

The substantial burden affecting over 280,000 adolescents nationally and persistence of high rates among 18-19 year-olds (50%) demonstrate that intensified multilevel efforts are essential to protect adolescent health, enable educational and economic opportunities, and break intergenerational poverty cycles.

Teenage pregnancy is not inevitable—it results from modifiable social, economic, and health system factors operating at multiple levels. With political commitment, adequate resources, evidence-based strategies accounting for both individual and community determinants, and sustained implementation, Cambodia can dramatically reduce teenage pregnancy, safeguarding the health, rights, and futures of its adolescent girls.

## ACKNOWLEDGMENTS

We acknowledge the National Institute of Statistics of Cambodia and ICF International for conducting the Cambodia Demographic and Health Survey and making the data publicly available for research. We thank all survey respondents for their participation and the field teams who collected these valuable data. We are grateful to the DHS Program for technical support and data access.

## FUNDING

This research received no specific grant from any funding agency in the public, commercial, or not-for-profit sectors. The CDHS 2021-22 was funded by the United States Agency for International Development (USAID), the Government of Cambodia, and other development partners.

## CONFLICTS OF INTEREST

The authors declare no conflicts of interest.

## DATA AVAILABILITY

The dataset analyzed in this study is publicly available from the DHS Program website (https://dhsprogram.com) upon registration and approval of a data request.

## Notes

### Competing Interest Statement

The authors have declared no competing interest.

### Funding Statement

This study did not receive any funding.

